# Rare variant analyses validate known ALS genes in a multi-ethnic population and identifies *ANTXR2* as a candidate in PLS

**DOI:** 10.1101/2023.09.30.23296353

**Authors:** Tess D. Pottinger, Joshua E. Motelow, Gundula Povysil, Cristiane A. Martins Moreno, Zhong Ren, Hemali Phatnani, The New York Genome Center ALS Sequencing Consortium, Timothy J. Aitman, Javier Santoyo-Lopez, Scottish Genomes Partnership, Hiroshi Mitsumoto, ALS COSMOS Study Group, PLS COSMOS Study Group, GTAC Investigators, David B. Goldstein, Matthew B. Harms

**Author notes:** **CORRESPONDING AUTHOR:** Matthew B. Harms, MD CUIMC/Neurological Institute of New York Department of Neurology, 710 West 168^208^ Street New York, NY 10032.

## Abstract

**Background:** Amyotrophic lateral sclerosis (ALS) is a neurodegenerative disease affecting over 30,000 people in the United States. It is characterized by the progressive decline of the nervous system that leads to the weakening of muscles which impacts physical function. Approximately, 15% of individuals diagnosed with ALS have a known genetic variant that contributes to their disease. As therapies that slow or prevent symptoms, such as antisense oligonucleotides, continue to develop, it is important to discover novel genes that could be targets for treatment. Additionally, as cohorts continue to grow, performing analyses in ALS subtypes, such as primary lateral sclerosis (PLS), becomes possible due to an increase in power. These analyses could highlight novel pathways in disease manifestation.

**Methods:** Building on our previous discoveries using rare variant association analyses, we conducted rare variant burden testing on a substantially larger cohort of 6,970 ALS patients from a large multi-ethnic cohort as well as 166 PLS patients, and 22,524 controls. We used intolerant domain percentiles based on sub-region Residual Variation Intolerance Score (subRVIS) that have been described previously in conjunction with gene based collapsing approaches to conduct burden testing to identify genes that associate with ALS and PLS.

**Results:** A gene based collapsing model showed significant associations with *SOD1*, *TARDBP*, and *TBK1* (OR=19.18, p = 3.67 × 10^−39^; OR=4.73, p = 2 × 10^−10^; OR=2.3, p = 7.49 × 10^−9^, respectively). These genes have been previously associated with ALS. Additionally, a significant novel control enriched gene, *ALKBH3* (p = 4.88 × 10^−7^), was protective for ALS in this model. An intolerant domain based collapsing model showed a significant improvement in identifying regions in *TARDBP* that associated with ALS (OR=10.08, p = 3.62 × 10^−16^). Our PLS protein truncating variant collapsing analysis demonstrated significant case enrichment in *ANTXR2* (p=8.38 × 10^−6^).

**Conclusions:** In a large multi-ethnic cohort of 6,970 ALS patients, rare variant burden testing validated known ALS genes and identified a novel potentially protective gene, *ALKBH3*. A first-ever analysis in 166 patients with PLS found a candidate association with loss-of-function mutations in *ANTXR2*.

## Background

Amyotrophic lateral sclerosis (ALS) is a rare neurodegenerative disease characterized by the progressive loss of upper motor neurons in the cortex and lower motor neurons of the brainstem and spinal cord. Even with FDA-approved disease modifying medication and palliation by artificial nutrition and ventilation, the prognosis is poor and death from accumulating paralysis occurs a median of 32 months after symptoms first manifest(1). Over the last 30 years, genetic study of the 5-10% of ALS patients with family history(2, 3) have securely implicated ∼20 monogenic causes and showed possible association to a similar number of genes (https://clinicalgenome.org/affiliation/40096/). Causative mutations in the most prevalent ALS genes (*C9ORF72, SOD1*, *TARDBP*, and *FUS)* explain ∼70% of familial ALS(4, 5). Due in part to incomplete penetrance, 10% of simplex ALS cases also carry mutations in these same genes(6).

A paucity of unsolved ALS pedigrees for family studies has intersected with falling sequencing costs for large-scale sequencing to allow gene discovery studies based on rare variant burden or collapsing methods on cohorts using predominantly simplex patients. Since our group first used this approach to implicate *TBK1* and *NEK1,* others have also identified *DNAJC7, TUBA4A* and several candidates(6–9). These analyses utilized the entire gene or recognizable functional domains as the regions for burden testing (6, 7) and were restricted to cohorts with European ancestry, or with less than 5% non-European ALS cases. Recognizing that power for discovery could be improved by a) increasing case and control numbers, b) diversifying the ancestries of participants, and c) collapsing on domains known to be intolerant of variation, we conducted both standard gene and intolerant domain-based collapsing analyses on 6,970 multi-ethnic ALS cases and ancestry-matched controls. Primary lateral sclerosis (PLS) is also a neurodegenerative disease of motor neurons with clinical features, neuropathology, and some genetics that overlaps with ALS(10–12). PLS is nearly always simplex and 20 times rarer than ALS(13). Because large-scale sequencing studies of ALS often include PLS patients, we were able to conduct a gene-based collapsing analysis in 166 PLS multi-ethnic cases and ancestry matched controls.

## Methods

### Study population

All samples and data came from participants that provided written, informed consent for genetic studies that had been IRB-approved at each contributing center. The study cohort includes participants from the Genomic Translation for ALS Care (GTAC study), the Columbia University Precision Medicine Initiative for ALS, the ALS COSMOS Study Group, the PLS COSMOS Study Group, the New York Genome Consortium, and the ALS Sequencing Consortium (IRB-approved genetic studies from Columbia University Medical Center, including the Coriell NINDS repository), University of Massachusetts at Worchester, Stanford University (including samples from Emory University School of Medicine, the Johns Hopkins University School of Medicine, and the University of California, San Diego), Massachusetts General Hospital Neurogenetics DNA Diagnostic Lab Repository, Duke University, McGill University (including contributions from Saint-Luc and Notre-Dame Hospital of the Centre Hospitalier de l’Université de Montréal [CHUM], [University of Montreal]), Gui de Chauliac Hospital of the CHU de Montpellier (Montpellier University), Pitié Salpêtrière Hospital, Fleurimont Hospital of the Centre Hospitalier Universitaire de Sherbrooke (CHUS) (University of Sherbrooke), Enfant Jésus Hospital of the Centre hospitalier affilié universitaire de Québec (CHA) (Laval University), Montreal General Hospital, Montreal Neurological Institute and Hospital of the McGill University Health Centre, the University of Edinburgh Scotland, and Washington University in St. Louis (including contributions from Houston Methodist Hospital, Virginia Mason Medical Center, University of Utah, and Cedars Sinai Medical Center). Participants were determined to have ALS or PLS by neuromuscular specialists at tertiary motor neuron disease care centers with expertise in distinguishing between the two. ALS diagnoses were based on the El Escorial Criteria in all cases. For PLS, explicit criteria requiring >3 years of symptoms without conversion to ALS were used for 79 of the 172 PLS participants (14). The criteria used for the remaining PLS diagnoses were not available.

Controls were selected from >100,000 whole-exome or -genome sequenced individuals housed in the IGM Data Repository. Individuals with known neurodegenerative disease were excluded. However, none of the controls were screened for neurodegenerative disease. All participants consented to the use of DNA in genetic research.

### Whole exome and genome sequencing

DNA sequencing was performed at Columbia University, the New York Genome Center, Duke University, McGill University, Stanford University, HudsonAlpha, and University of Massachusetts, Worcester. Kits used to conduct whole-exome capture are as follows: Agilent All Exon kits (50MB, 65MB, and CRE), Nimblegen SeqCap EZ Exome Enrichment kits (V2.0, V3.0, VCRome, and MedExome), and IDT Exome Enrichment panel. There were 2,185 participants with ALS who were sequenced using Nimblegen SeqCap EZ Exome Enrichment kits and 51 who were sequenced using the IDT Exome Enrichment panel (**Supplemental Table 1**). While 1,272 controls were evaluated using the Aligent All Exon kits, 8,498 with the IDT Exome Enrichment panel, and 11,201 with the Nimblegen SeqCap EZ Exome Enrichment kits. Sequencing was performed using Illumina GAIIx, HiSeq 2000, HiSeq 2500, and NovaSeq 6000 sequencers according to standard protocols. Whole genome sequencing was conducted at the New York Genome Center and in-house at the IGM. Sample-level BAM files were transferred from the New York Genome Center to the IGM (n = 3,418). An additional 1,316 genomes were processed by the IGM. There were 1,553 genomes in our control cohort (**Supplemental Table 1**). Data were aligned to the human reference genome (NCBI Build 37) using DRAGEN (Edico Genome, San Diego, CA, USA). Picard (http://picard.sourceforge.net) was used to remove duplicate reads and to process lane-level BAM files to create a sample-level BAM file. GATK was used to recalibrate base quality scores, realign around indels, and call variants utilizing the Best Practices recommendations v3.6 (15). Variants were annotated using ClinEff and the Analysis Tool for Annotated Variants (ATAV), an in-house IGM annotation tool (16). Variants were annotated with the Genome Aggregation Database (gnomAD) v2.1 frequencies, regional-intolerance metrics, and the clinical annotations by the Human Gene Mutation Database (HGMD), ClinVar, and Online Mendelian Inheritance in Man (OMIM). Exonic regions were retained for downstream statistical analyses.

### Sample and variant quality control

Samples reporting >2% contamination according to verifyBamID (17) and those with consensus coding sequence (CCDS release 20) <90% were excluded from these analyses. KING (18) was used to test for relatedness. Only unrelated (up to second-degree) individuals were included in these analyses. For related pairs, samples were chosen to prefer cases. Samples where X:Y coverage ratios did not match expected sex were excluded.

Only variants within the CCDS or the 2 bp canonical sites were included in these analyses. These variants were also required to have a quality score of at least 50, a quality by depth score of at least 5, genotype quality score of at least 20, read position rank sum of at least −3, mapping quality score of at least 40, mapping quality rank sum greater than −10, and a minimum coverage of at least 10. SNVs had a maximum Fisher’s strand bias of 60, while indels had a maximum of 200. For heterozygous genotypes, the alternative allele ratio was required to be greater than or equal to 30%. Only variants with the GATK Variant Quality Score Recalibration filter “PASS”, “VQSRTrancheSNP90.00to99.00”, or “VQSRTrancheSNP99.00to99.90” were included. Variants were excluded if they were marked by EVS, ExAC, or gnomAD as being failures (http://evs.gs.washington.edu/EVS/).

### Clustering, ancestry, and coverage harmonization

A neural network pre-trained on samples of known ancestry was used to calculate probability estimates for six ancestry groups (African, East Asian, European, Hispanic, Middle Eastern, and South Asian). Methods for characterizing samples into clusters has been previously described (19).

To ensure balanced sequencing coverage of evaluated sites between cases and controls, we imposed a statistical test of independence between the case/control status and coverage as previously described (20). Sites were removed where the absolute difference in percentages of cases and controls with at least 10x coverage was greater than 7%. Samples were then pruned using this method on a cluster-by-cluster basis. Through this approach, approximately 7-11% were removed. Clusters with less than 5 participants were not included in these analyses, thereby removing 6 participants with PLS but none with ALS.

### Variant-level statistical analysis

The models that were used to test for associations of nonsynonymous coding or canonical splice variants with outcome included variants with MAF <0.1% for each population represented in gnomad and internal AF of <0.1%. Models tested were a standard gene-unit collapsing analysis, and a domain-unit analysis. The models used for these analyses were previously described (7). A domain-based approach utilizing sub-region Residual Variation Intolerance Score (subRVIS) domain percentage(7) with a threshold of 25 was also used to evaluate case enrichment of rare variants. The full list of 18,653 CCDS genes was analyzed for each model.

Genes with at least one qualifying variant were included for analyses. As we are meta analyzing across clusters an exact 2-sided Cochran-Mantel-Haenszel test was used (using the statistical package in R v3.6). Study-wide significance was determined by accounting for 6 nonsynonymous models-multiplicity-adjusted significance threshold α = 4.9 × 10^−7^ (**Supplemental Table 2**). Model inflation was calculated using empirical (permutation-based) expected probability distributions as described by Povysil and colleagues (19).

### ALS and PLS rare variant burden testing

We conducted both standard gene and intolerant domain-based collapsing analyses on 6,970 multi-ethnic ALS cases (87% European) and 22,534 ancestry-matched controls. Standard gene collapsing analyses identified case enrichment of rare variants (minor allele frequency of 0.001) in an ALS cohort with 12 sub-population groups (**Supplemental Figure 1A**) that correspond to ancestry-based clusters (**Supplemental Figure 1B; Supplemental Table 3**). Analyses were conducted on clusters with at least 3 cases. Controls were drawn from individuals sequenced for phenotypes/diseases with no known association with ALS (**Supplemental Table 4**). As expected, a negative control analysis for rare synonymous variants found no case-enrichment (**Supplemental Figure 2**). Because gene-based collapsing considers variation across the entire gene, regions that are tolerant of variation could swamp case-enrichment signals originating from regions that are intolerant of variation (7). To overcome this limitation, we conducted rare variant collapsing on domains that are intolerant to variation as defined as a subRVIS domain score threshold of 25, a cutoff based on threshold testing.

As large-scale sequencing studies of ALS often include PLS patients, we were able to conduct a gene-based collapsing analysis in 166 PLS multi-ethnic cases (88% European) and 17,695 ancestry matched controls (**Supplemental Figure 3; Supplemental Tables 5**). We expected the study would be underpowered for securely implicating causative genes but used this as an opportunity to generate candidates for future study.

### ALS gene set enrichment analyses

An ALS gene set enrichment analysis was conducted using the gene strength association list outlined in **Table 1**. We utilized the qualifying variants that were associated with ALS in each gene set category and used the exact two-sided CMH test to analyze burden of ALS genes defined by gene set. These lists were curated using data published by Gregory and colleagues (21). As outlined, “ALS Confirmed” genes were found to have ample published replication evidence, while ‘ALS Plus’ genes had some replication data and/or functional evidence for an association with ALS. However, ‘ALS Replication Needed’ genes, required additional replication analyses and/or functional data, and ‘ALS Weak Evidence’ genes were genes that overlapped with ALS phenotypically.

**Table 1.**
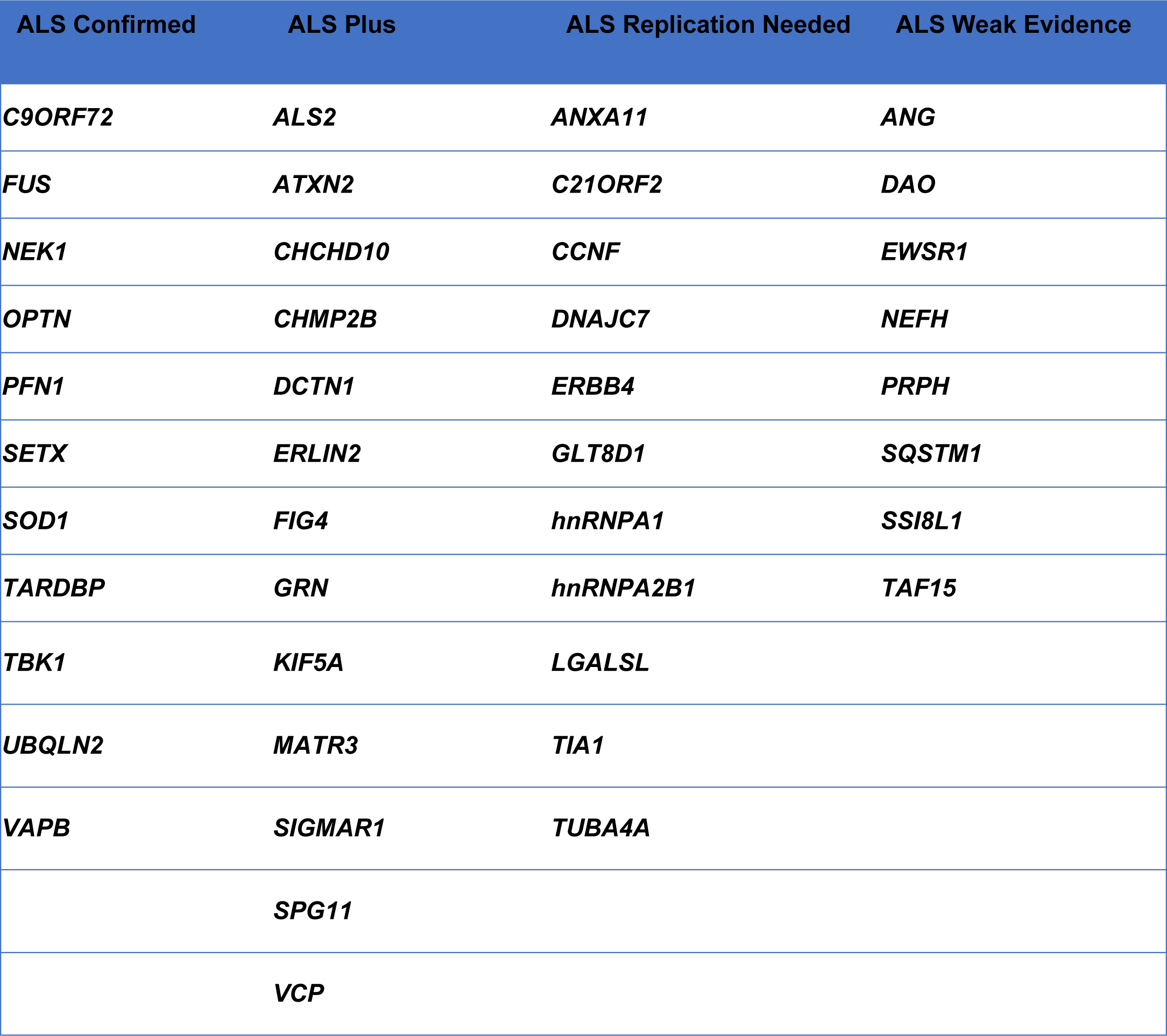
ALS gene association strength.

## Results

### Rare variant burden testing

Collapsing analysis of all rare functional variants (missense and protein truncating variants) (**Supplemental Table 2**) found genome-wide and study-wide significant (p < 4.9 × 10^−7^) case-enrichment for known ALS genes *SOD1*, *TARDBP*, *TBK1* (OR=19.18, p = 3.67 × 10^−39^; OR=4.73, p = 2 × 10^−10^; OR=2.3, p = 7.49 × 10^−9^, respectively) and control-enrichment for *ALKBH3* (OR=0.26, p = 4.88 × 10^−7^) (**Figure 1A; Supplemental Data**). Although *SOD1, TBK1* and *TARDBP* are definitive ALS genes, we were intrigued by the identification of control-enriched *ALKBH3.* Control-enrichment was not explained by sequencing methodology, ancestry cluster, or specific phenotype/disease population within the control cohort. Because *ALKBH3* plays a role in DNA repair(22), a mechanism increasingly implicated in ALS pathogenesis(23), we attempted to replicate this novel association by analyzing summary statistics from the Project MinE cohort, which is similar in size to ours (24). None of the available models focused on variation as rare as in our analyses, but at a higher minor allele frequency (MAF) for qualifying variants (0.005), a minor degree of control-enrichment was in fact observed (OR= 0.56, p = 3.96 × 10^−4^). This raises the possibility that rare missense and protein truncating variants (PTVs) in *ALKBH3* could protect from ALS, a finding that requires validation in large cohorts.

**Figure 1.**
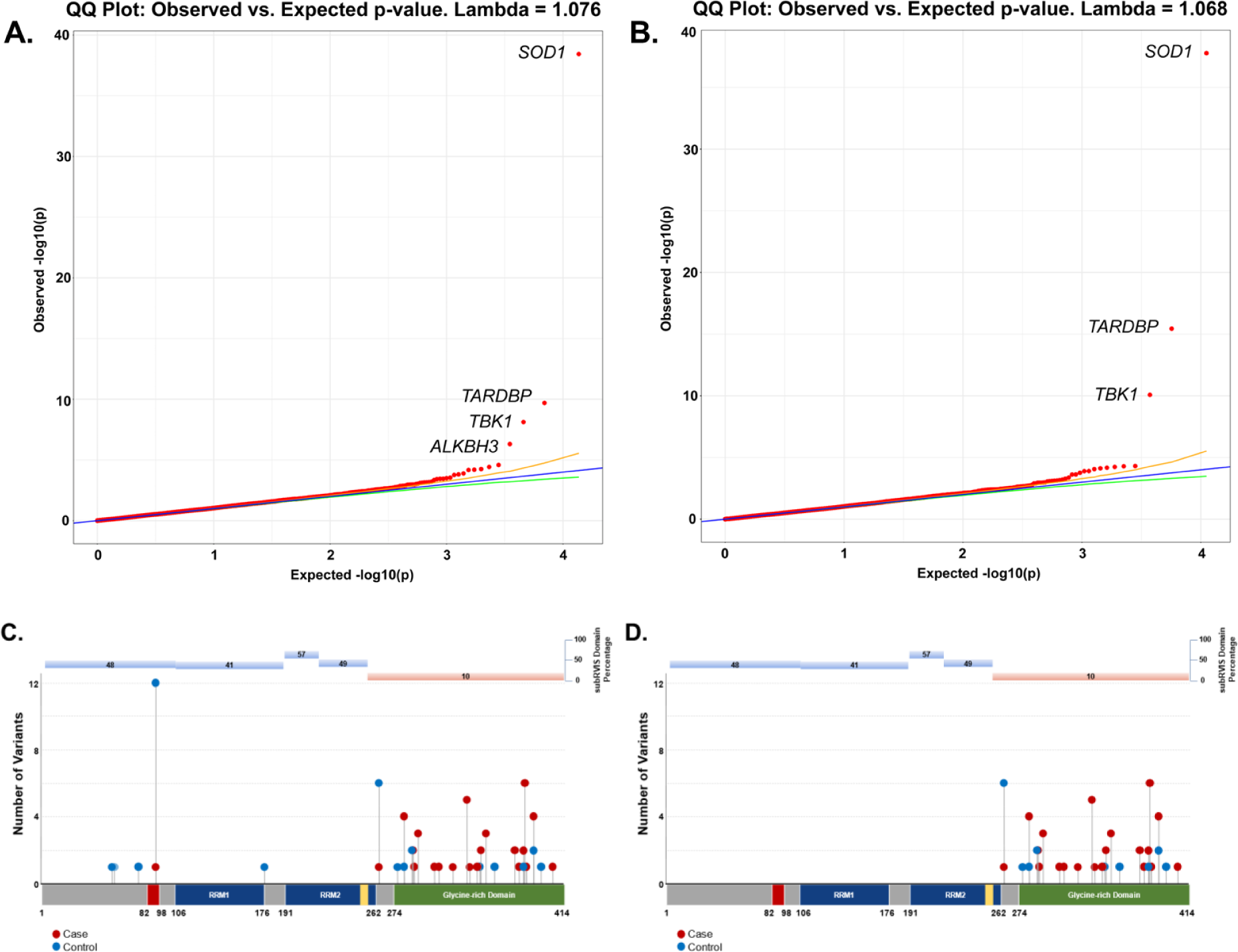
Q-Q plots of gene- and domain-level collapsing of ALL functional coding variants in ALS cohort. (A) The results for a standard gene-level collapsing of 6,970 ALS cases and 22,524 controls. P-values were generated using an exact two-sided Cochran-Mantel-Haenszel (CMH) by gene by cluster. The genes with the top associations that achieved study-wide significance of p<4.9×10^−7^ (*SOD1* (OR=19.18)*, TARDBP* (OR=4.73)*, TBK1* (OR=2.3), and *ALKBH3* (OR=0.26)) are labeled. *SOD1, TARDBP, TBK1* have been previously implicated in rare variant association studies of ALS. Yellow and green lines indicate the 2.5^th^ and 97.5^th^ percentile of expected p-values, respectively. (B) The results for the domain-based collapsing restricting qualifying variants to those with subRVIS domain percentage score < 25 of 6,970 cases and 22,524 controls. P-values were generated using an exact two-sided Cochran-Mantel-Haenszel (CMH) by gene by cluster. The genes with the top associations (*SOD1* (OR=20.63)*, TARDBP* (OR=10.08), and *TBK1* (OR=3.15)) are labeled. (C) Standard gene-level collapsing model showed 44 qualifying variants in cases (red circles) and 31 in controls (blue circles) for *TARDBP* (D) subRVIS domain collapsing improved association by removing control variants (cases = 43; controls = 15). Regions with subRVIS domain percentage below 25 are highlighted in orange while those above this threshold are highlighted in blue. A one tailed z-score showed that there were significantly less controls in the intolerant domain as indicated by subRVIS domain percentage score < 25 (p=0.031).

Intolerant domain analyses implicated the same three known ALS genes (*SOD1*, *TARDBP*, and *TBK1* at OR=20.63, p = 1.68 × 10^−38^; OR=10.08, p = 3.62 × 10^−16^; and OR=3.15, p = 8.38 × 10^−11^, respectively) (**Figure 1B; Supplemental Data**). The intolerant domain analysis did not improve over the gene-based analysis for *SOD1* or *TBK1* (**Figure 2**; **Figure 3**) but doubled the odds ratio and significantly lowered the p-value obtained for *TARDBP.* The improvement of the intolerant domain model (**Figure 1C, 1D**) stemmed from a significant drop (one-tailed z-score p=0.031) in the number of qualifying variants found in controls dispersed across tolerant regions, while highlighting qualifying variants in ALS cases predominantly in the intolerant C-terminal region.

**Figure 2.**
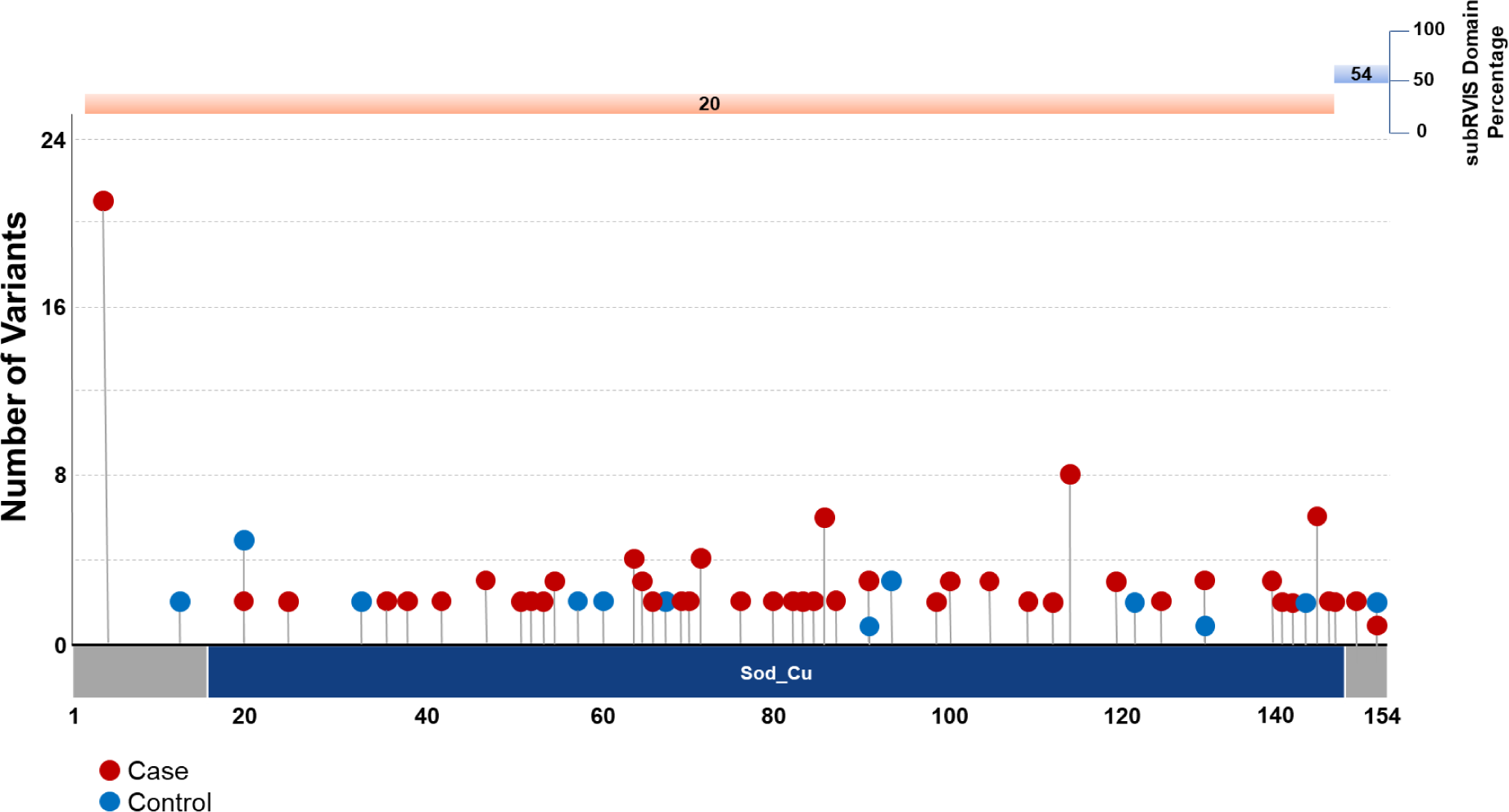
Plot of gene- and domain-level collapsing of ALL *SOD1* functional coding variants. Standard gene-level collapsing model showed 93 qualifying variants in cases (red circles) and 18 in controls (blue circles) for *SOD1*. subRVIS domain collapsing improved association by removing control variants (cases = 90; controls = 16). Regions with subRVIS domain percentage below 25 are highlighted in orange while those above this threshold are highlighted in blue. However, a one tailed z-score showed that the differences in the number of controls in the intolerant domain was not significantly lower than those in the entire gene as indicated by subRVIS domain percentage score < 25 (p=0.4).

**Figure 3.**
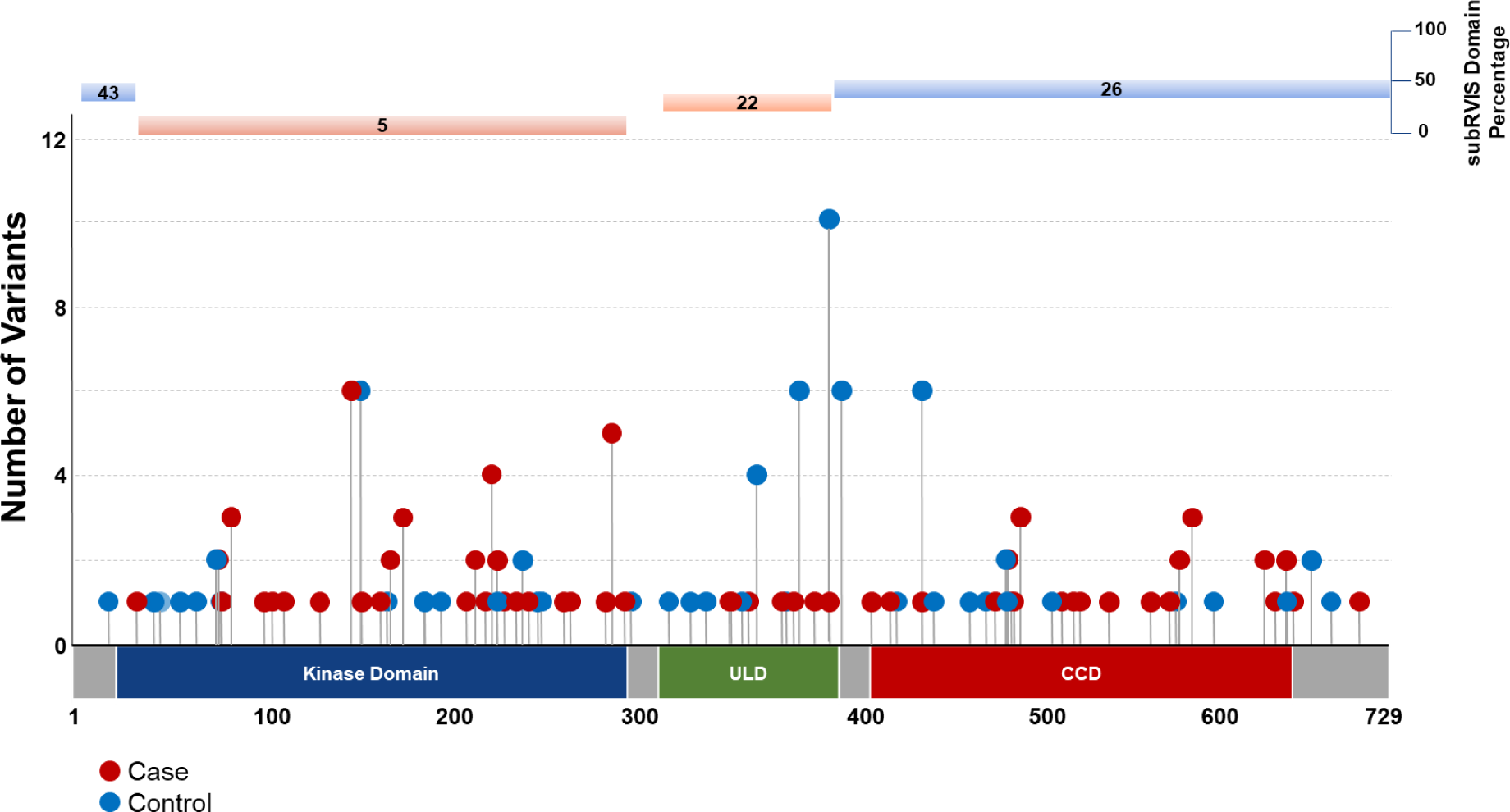
Plot of gene- and domain-level collapsing of ALL *TBK1* functional coding variants. Standard gene-level collapsing model showed 73 qualifying variants in cases (red circles) and 143 in controls (blue circles) for *TBK1*. subRVIS domain collapsing improved association by removing control variants (cases = 47; controls = 72). Regions with subRVIS domain percentage below 25 are highlighted in orange while those above this threshold are highlighted in blue. However, a one tailed z-score showed that the differences in the number of controls in the intolerant domain was not significantly lower than those in the entire gene as indicated by subRVIS domain percentage score < 25 (p=0.3).

Although most models showed no significant genes, the dominant PTV model showed significant case enrichment for *ANTXR2* (OR=174.57, p=8.38 × 10^−6^) (**Figure 4; Supplemental Table 6; Supplemental Data**), a gene associated with brain connectivity changes and Alzheimer’s disease(25). Currently, there are no additional large sequencing studies of PLS in which we could attempt replication.

**Figure 4.**
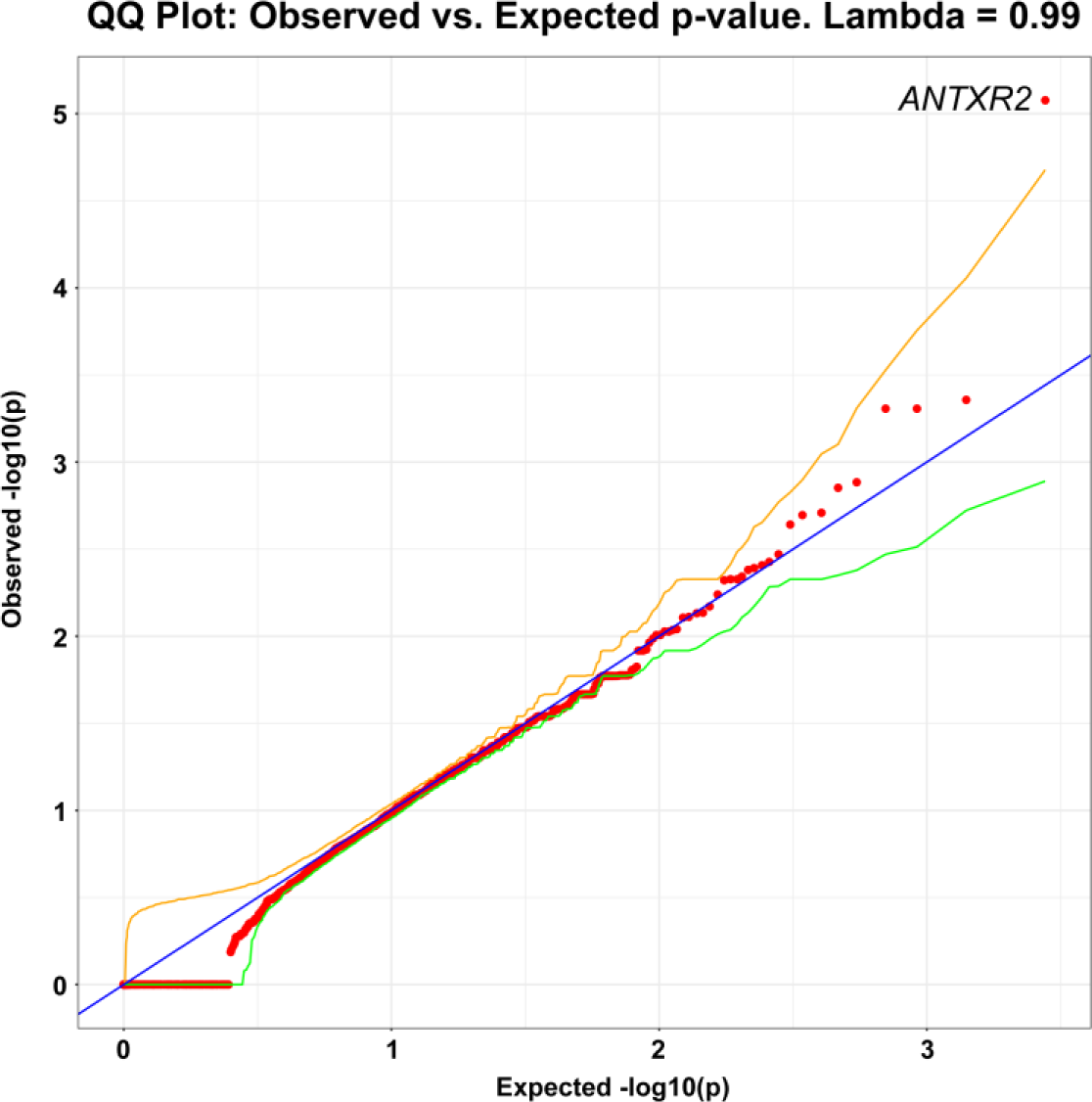
Q-Q plot of gene-level collapsing of protein truncating variants (PTV) in PLS cohort. The results for a standard gene-level collapsing of 166 PLS cases and 17,695 controls. P-values were generated using an exact two-sided Cochran-Mantel-Haenszel (CMH) by gene by cluster. The gene with the top associations that achieved genome-wide significance of p<8.38×10^−6^ (*ANTXR2* (OR=174.57)) is labeled. *ANTXR2* has not been previously implicated in rare variant association studies of PLS. Yellow and green lines indicate the 2.5^th^ and 97.5^th^ percentile of expected p-values, respectively.

### ALS gene set enrichment analyses

A gene set enrichment analysis of genes that were defined as ‘ALS Confirmed’ were significantly associated with the ALS for all dominant models, including PTV only (p = 9.12 × 10^−24^), Missense & PTV (p = 6.63 × 10^−19^), and Missense only (p = 1.03 × 10^−19^) (**Figure 5**). The synonymous model, which served as a control, showed no association (p = 0.79) between these genes and ALS. Genes that are weakly associated with ALS, ‘ALS Weak Evidence’, showed no significant enrichment of rare variants for the 4 models that were analyzed. The group of genes that were described as needing additional replication studies, ‘ALS Replication Needed’, showed a significant association with rare variants and ALS for the Missense & PTV model (p = 4.6 × 10^−3^). For all other models, rare variants in these genes were not significantly associated with ALS. An analysis of genes that are characterized as ‘ALS Plus’ showed no significant association of rare variants with ALS for the 4 models that were analyzed.

**Figure 5.**
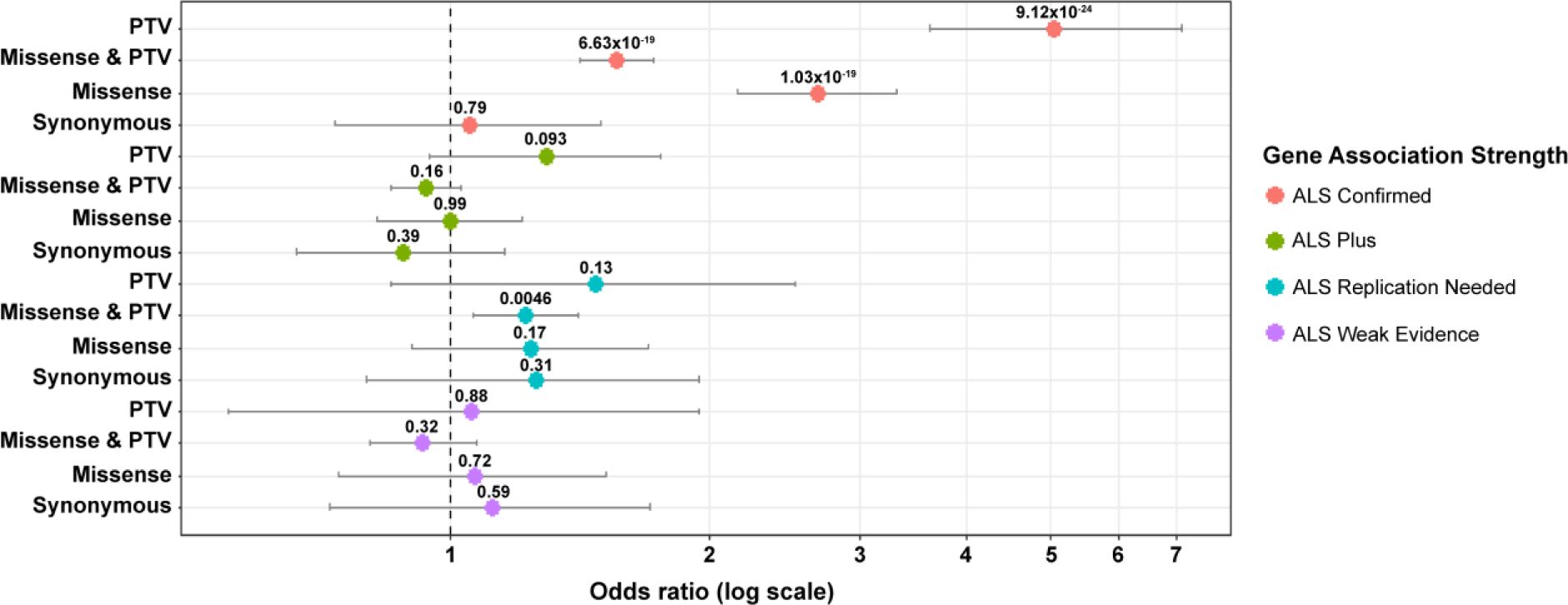
Forest plot of ALS genes by model. Rare variants in “ALS Confirmed” genes were significantly associated with ALS in all gene-based collapsing models except the control synonymous model. Rare variants in “ALS Plus” genes were associated with ALS in “Missense & PTV” gene-based collapsing model. There was no association with ALS of rare variants in “ALS Replication Needed” and “ALS Weak Evidence” genes. Pooled odds ratio, 95% confidence intervals, and p-values were generated from exact two-sided Cochran-Mantel-Haenszel (CMH) tests.

## Discussion

### Burden testing

Conducting genic and intolerant domain based rare variant burden testing in a large multi-ethnic population provides insight into novel and established biological mechanisms in disease manifestations. Additionally, analyzing specific disease subtypes can capture critical disease pathways that could be targets for clinical intervention. Here we show, that performing burden testing in multi-ethnic populations and in disease subtypes found novel genetic associations in individuals diagnosed with ALS and PLS. These analyses implicated ALS genes that have previously been identified (*SOD1*, *TARDBP*, and *TBK1*). We also identified *ALKBH3* as a potentially protective gene that warrants further study in additional cohorts. In addition, we conducted the first rare variant collapsing analysis in PLS, identifying PTVs in *ANTXR2*. This gene will need to be investigated further in larger PLS cohorts or in targeted functional analyses. Lastly, gene set enrichment analyses provide evidence that genes known to be associated with ALS show strong evidence to have a rare variant burden especially for protein truncating variants.

### ALKBH3 associates with ALS

We found that genic burden testing of individuals diagnosed with ALS identified known risk genes (*SOD1*, *TARDBP*, and *TBK1*) and a novel protective gene (*ALKBH3*). *ALKBH3* encodes for AlkB homolog 3, Alpha-Ketoglutarate Dependent Dioxygenase which protects against the cytotoxicity of methylating agents by repair of the specific DNA lesions (26–28). ALKBH3 potentially acts as a putative hyperactive promotor to suppress transcription associated DNA damage of highly expressed genes (29). Genes that play a role in DNA repair and DNA damage response such as *TARDBP*, *FUS*, and *NEK1* (30–33) are known to play a role in ALS potentially through neuronal death pathways.

### ANTXR2 associated with PLS

Genic burden testing of protein truncating variants on individuals with PLS identified a suggestive gene (*ANTXR2*). *ANTXR2* encodes a receptor for anthrax toxin that may be involved in extracellular matrix adhesion. Variants in this gene have been associated with hyaline fibromatosis (34, 35), and has been shown to play a role in angiogenesis (36). This finding adds to the number angiogenic genes that have been implicated in ALS including *VEGF* and *ANG* (37).

While we identified a potentially important gene that is associated with PLS, we were limited in our sample size and will therefore need additional cohorts or functional studies to further investigate this finding. Additionally, there are potentially more ALS subtypes that could be investigated to better understand this heterogeneous disease. Lastly, unknown confounders could be contributing to the signal that are found in these association analyses.

## Conclusions

In summary, we performed the largest rare variant analyses of a multi-ethnic population of patients with ALS to date. Our analysis did not identify new ALS risk genes but demonstrated that collapsing models informed by regions of intolerance can be useful for identifying genes where disease-associated variation is limited to regions with low background variation. This analysis also confirmed the association of the C-terminal domain of *TARDBP.* We also identified *ALKBH3* as a potentially protective gene that warrants further study in additional and larger cohorts. Finally, we conducted the first rare variant collapsing analysis in PLS, identifying PTVs in *ANTXR2* as a candidate disease gene. This association and potential mechanisms for PTVs in this gene will need to be investigated further in larger PLS cohorts.

It is important to note that this analysis doubled the number of ALS cases and quadrupled the number of controls from our first study(6) but remained underpowered for the identification of new ALS genes. A recently published rare variant burden analysis with a similar number of ALS cases did not identify new genes(24) either, emphasizing the need for increasingly large genomically characterized ALS cohorts, especially in non-European populations.

## Data Availability

All summary data produced in the present work are contained in the manuscript

## ACKNOWLEDGEMENTS

We thank the following groups for contributing ALS/PLS samples, sequencing, or clinical data:

## New York Genome Center ALS Consortium

J. Kwan, D. Sareen, J.R. Broach, Z. Simmons, X. Arcila-Londono, E.B. Lee, V.M. Van Deerlin, E. Fraenkel, L.W. Ostrow, F. Baas, N. Zaitlen, J.D. Berry, A. Malaspina, P. Fratta, G.A. Cox, L.M. Thompson, S. Finkbeiner, E. Dardiotis, T.M. Miller, S. Chandran, S. Pal, E. Hornstein, D.J. MacGowan, T. Heiman-Patterson, M.G. Hammell, N.A. Patsopoulos, J. Dubnau, and A. Nath.

## ALS Exome Sequencing Consortium

S.H. Appel, R.H. Baloh, R.S. Bedlack, R. Brown, W.K. Chung, S. Gibson, J.D. Glass, A. Gitler, D.B. Goldstein, T.M. Miller, R.M. Myers, S.M. Pulst, J.M. Ravits, G. Rouleau, E. Greene, N. Shneider, and W.W. Xin;

## Genomic Translation for ALS Care (GTAC) study

S.H. Appel, R.H. Baloh, R.S. Bedlack, S. Chandran, L. Foster, S. Goutman, E. Green, C. Karam, D. Lacomis, G. Manousakis, T.M. Miller, S. Pal, D. Sareen, A. Sherman, Z. Simmons, L. Wang.

## ALS COSMOS Study Sites Group

**Columbia University Coordinating Center, NY, NY:** Hiroshi Mitsumoto, MD, DSc, Pam Factor-Litvak, PhD, Regina Santella, PhD, Howard Andrews, PHD; **Texas Neurology, P.A., Dallas, TX:** Daragh Heitzman, MD; **Duke University, Durham, NC:** Richard S. Bedlack, MD, PhD; **California Pacific Medical Center, San Francisco, CA:** Jonathan S. Katz, MD, Robert Miller, MD, Dallas Forshew; **University of Kansas, Kansas City, KS:** Richard J. Barohn, MD, PhD; **Mayo Clinic, Rochester, MN; D**r. Eric J. Sorenson, MD; **University of California - Davis, Sacramento, CA:** Bjorn E. Oskarsson, MD, PhD; **University of Kentucky, Lexington, KY:** Edward J. Kasarskis, MD, PhD; **University of California - San Francisco, San Francisco, CA:** Catherine Lomen-Hoerth, MD, PhD, Jennifer Murphy, PhD; **University of Colorado, Aurora, CO:** Yvonne D. Rollins, MD, PhD; **University of California – Irvine, Orange, CA:** Tahseen Mozaffar, MD; **University of Nebraska, Omaha, NE;** J. Americo M. Fernandes, MD; **University of Iowa, Iowa City, IA:** Andrea J. Swenson, MD; **University of Texas - Southwestern, Dallas:** Sharon P. Nations, MD; **SUNY - Upstate Medical University, Syracuse, NY:** Jeremy M. Shefner, MD, PhD; and **Hospital for Special Care, New Britain, CT:** Jinsy A. Andrews, MD, MS, Dr. Agnes Koczon-Jaremko.

## PLS COSMOS Study Group

**Columbia University Irving Medical Center, NY, NY:** Hiroshi Mitsumoto, MD, DSc, Peter L. Nagy, MD, PhD, Pam Factor-Litvak, PhD, PhD, Rejina Santella, PhD, Howard Andrews, PhD, Raymond Goetz, PhD; **Icahn School of Medicine - Mount Sinai, NY, NY:** Chris Gennings, PhD; **University of California - San Francisco, San Fransisco, CA:** Jennifer Murphy, PhD; **National Institute of Neurological Disorders and Stroke, Bethesda, MD:** Mary Kay Floeter, MD, PhD; **University of Kansan Medical Center, Kansas City, KS:** Richard J. Barohn, MD; **University of Texas, Dallus, TX:** Sharon Nations, MD; **Western University, London, Ontario**: Christen Shoesmith, MD; and **University of Kentucky, Louisville, KT**: Edward Kasarskis, MD, PhD.

We thank The Washington Heights–Inwood Columbia Aging Project (WHICAP) for the contribution of control samples. We also thank the WHICAP study participants and the WHICAP research and support staff for their contributions to this study: K. Welsh-Bomer, C. Hulette, and J. Burke; D. Valle, J. Hoover-Fong, and N. Sobriera; A. Poduri; S. Palmer; R. Buckley; K. Newby; The Murdock Study Community Registry and Biorepository Pro00011196; National Institute of Allergy and Infectious Diseases Center for HIV/AIDS Vaccine Immunology (CHAVI) (U19-AI067854); National Institute of Allergy and Infectious Diseases Center for HIV/AIDS Vaccine Immunology and Immunogen Discovery (UM1-AI100645); CHAVI Funding; R. Ottman; V. Shashi; S. Berkovic, I. Scheffer, and B. Grinton; The Epi4K Consortium and Epilepsy Phenome/Genome Project; C. Depondt, S. Sisodiya, G. Cavalleri, and N. Delanty; S. Hirose; C. Woods, C. Village, K. Schmader, S. McDonald, M. Yanamadala, and H. White; G. Nestadt, J. Samuels, and Y. Wang; D. Levy; E. Pras, D. Lancet, and Z. Farfel; S. Chen; R. Wapner; C. Moylan, A. Mae Diehl, and M. Abdelmalek; DUHS (Duke University Health System) Nonalcoholic Fatty Liver Disease Research Database and Specimen Repository; M. Winn and R. Gbadegesin; M. Hauser; S. Delaney; A. Need and J. McEvoy; M. Walker; M. Sum; Undiagnosed Diseases Network; National Institute on Aging (R01AG037212, P01AG007232).

## Supplemental Figures and Tables

**Supplemental Figure 1.**
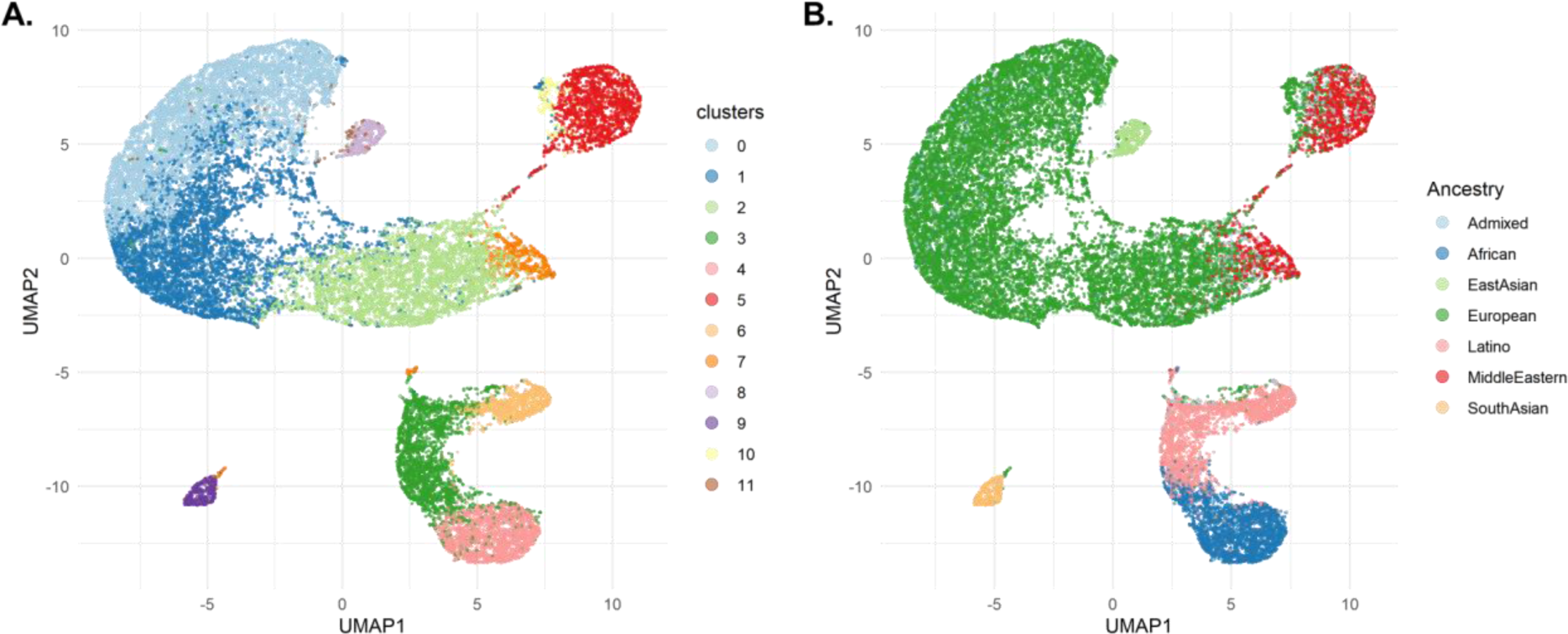
UMAP plot of ALS participants. (A) Cluster assignment (B) Predicted ancestry

**Supplemental Figure 2.**
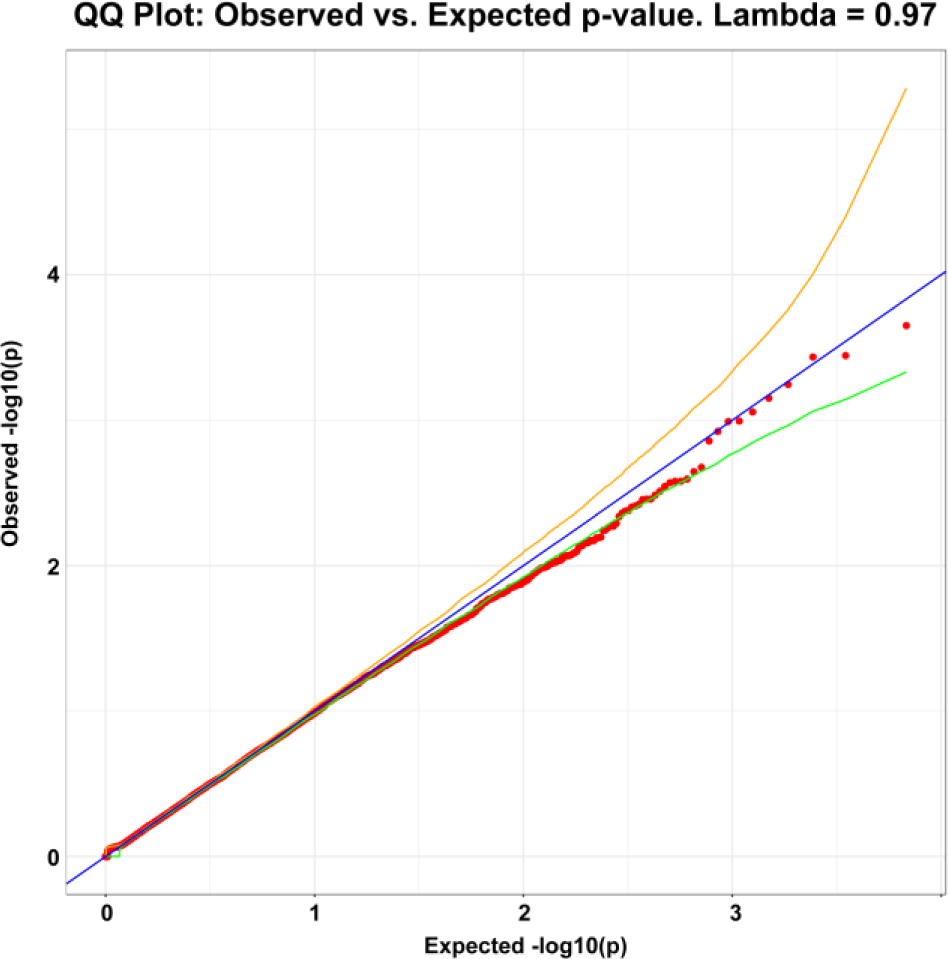
Q-Q plots of gene level collapsing in a Synonymous model. Yellow and green lines indicate the 2.5^th^ and 97.5^th^ percentile of expected p-values, respectively. The genomic inflation factor, lambda (λ), is 0.97 indicating no inflation. We generated p values from the exact two-sided Cochran-Mantel-Haenszel (CMH) test by gene by cluster to indicate a different carrier status of affected individuals in comparison to control individuals.

**Supplemental Figure 3.**
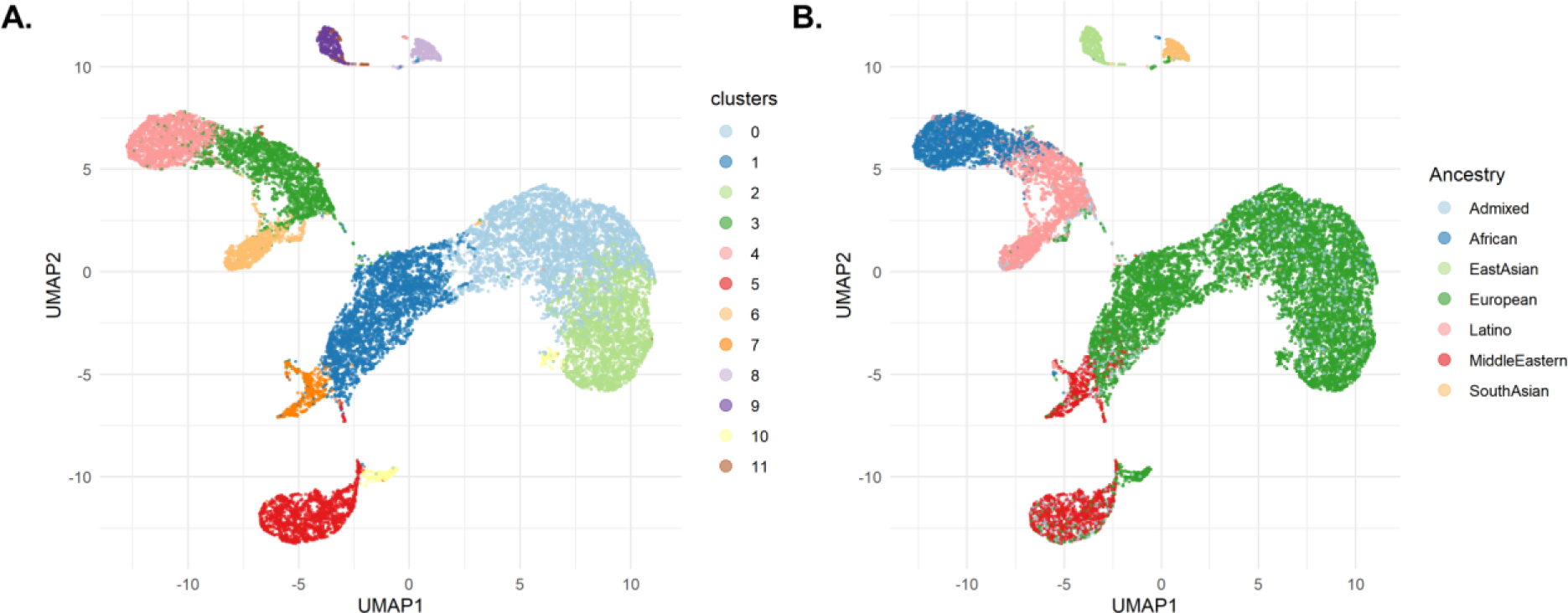
UMAP plot of PLS participants. (A) Cluster assignment (B) Predicted ancestry

**Supplemental Table 1.**
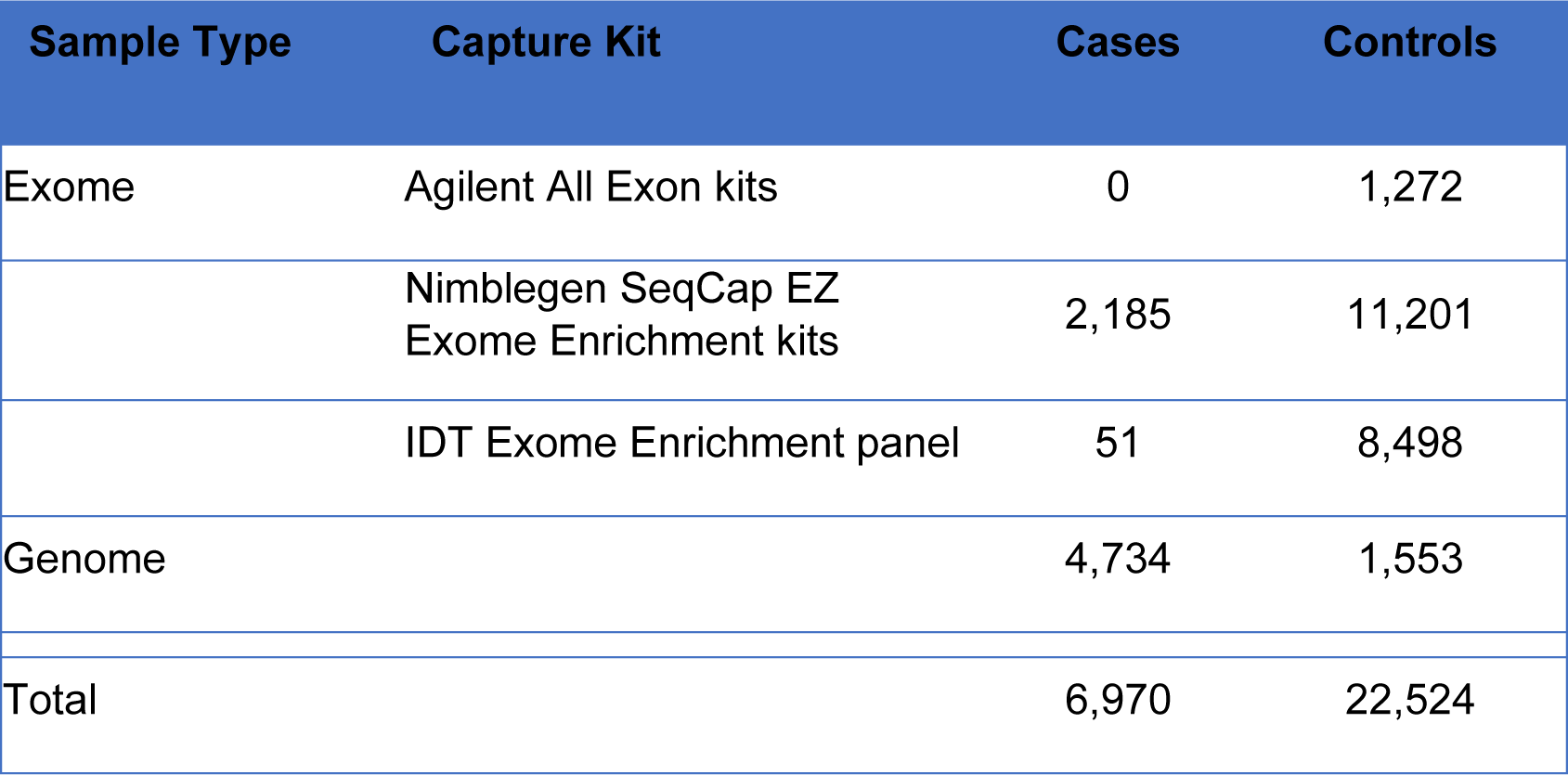
Sequencing kits used for cohort.

| Sample Type | Capture Kit | Cases | Controls |
| --- | --- | --- | --- |
| Exome | Agilent All Exon kits | 0 | 1,272 |
|  | Nimblegen SeqCap EZ Exome Enrichment kits | 2,185 | 11,201 |
|  | IDT Exome Enrichment panel | 51 | 8,498 |
| Genome |  | 4,734 | 1,553 |
| Total |  | 6,970 | 22,524 |

**Supplemental Table 2.**
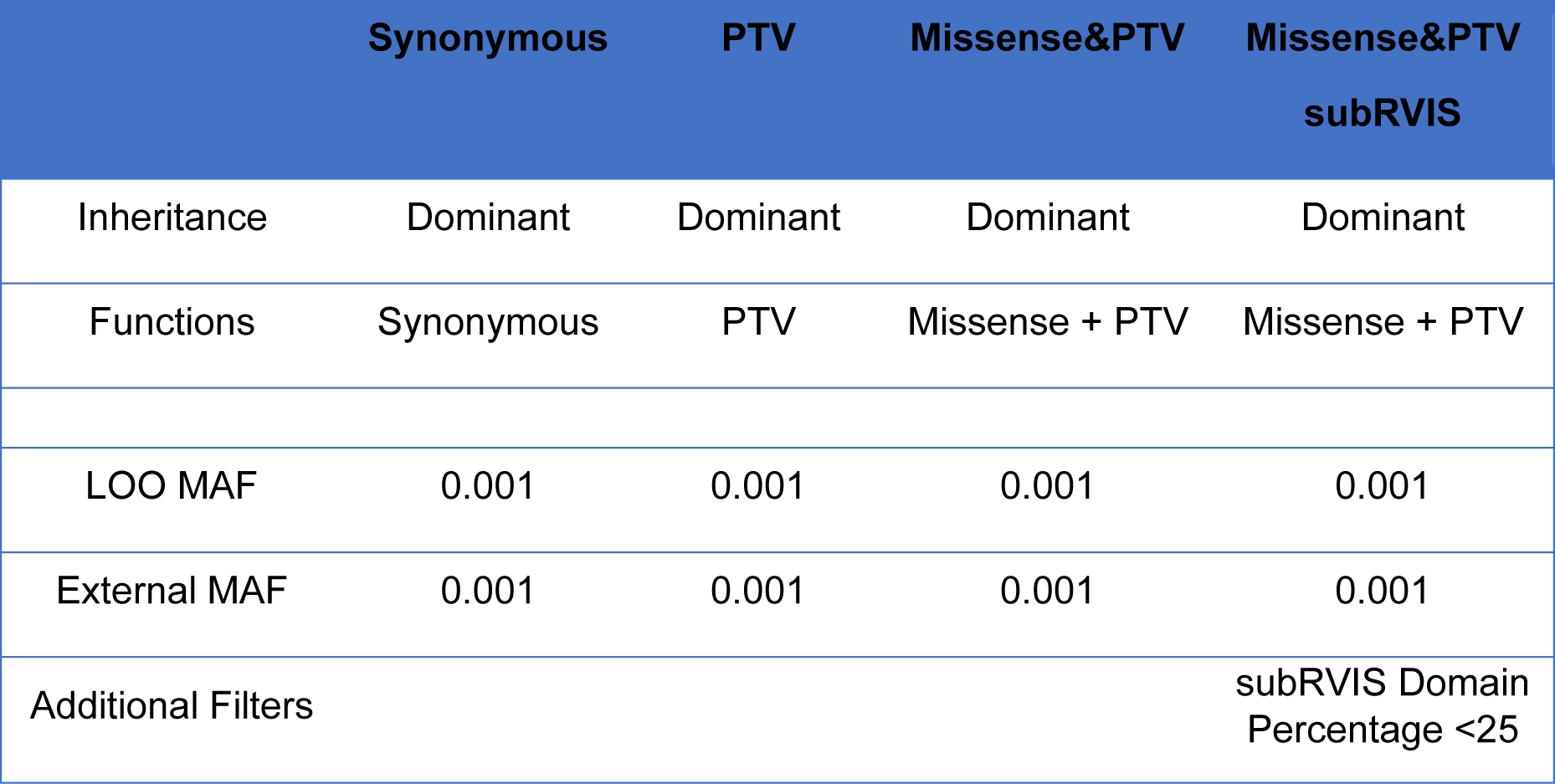
Description of collapsing models.

**Supplemental Table 3.** Cluster sizes for ALS cohort.

| Cluster | Ancestry | Cases | Controls | Ratio |
| --- | --- | --- | --- | --- |
| 0 | European1 | 2,620 | 4,792 | 0.55 |
| 1 | European2 | 2,502 | 3,989 | 0.63 |
| 2 | European3 | 922 | 3,868 | 0.24 |
| 3 | Latino1 | 104 | 2,788 | 0.037 |
| 4 | African1 | 204 | 2,102 | 0.097 |
| 5 | MiddleEastern1 | 294 | 1,924 | 0.15 |
| 6 | Latino2 | 110 | 1,068 | 0.1 |
| 7 | MiddleEastern2 | 86 | 669 | 0.13 |
| 8 | EastAsian1 | 35 | 496 | 0.071 |
| 9 | SouthAsian1 | 40 | 457 | 0.088 |
| 10 | European4 | 36 | 265 | 0.14 |
| 11 | Admixed1 | 17 | 106 | 0.15 |
| Total |  | 6,970 | 22,524 | 0.31 |

**Supplemental Table 4.** Phenotypes of control participants.

| Control Phenotype | Number of Participants |
| --- | --- |
| Kidney and urological disease | 8,416 (37.36%) |
| Healthy family member of a different study | 6,297 (27.96%) |
| Control of different study | 3,156 (14.01%) |
| Epilepsy | 2,892 (12.84%) |
| Obsessive compulsive disorder | 721 (3.2%) |
| Pulmonary disease | 403 (1.79%) |
| Hematological disease | 256 (1.13%) |
| Schizophrenia | 161 (0.71%) |
| Infectious disease | 107 (0.48%) |
| Liver disease | 82 (0.36%) |
| Primary immune deficiency | 33 (0.15%) |
| Total | 22,524 |

**Supplemental Table 5.** Cluster sizes for PLS cohort.

| Cluster | Ancestry | Cases | Controls | Ratio |
| --- | --- | --- | --- | --- |
| 0 | European1 | 90 | 4,792 | 0.017 |
| 1 | European2 | 19 | 3,989 | 0.0047 |
| 2 | European3 | 41 | 3,868 | 0.012 |
| 3 | Latino1 | 1 | 2,788 | 0.00039 |
| 4 | African1 | 3 | 2,102 | 0.0013 |
| 5 | MiddleEastern1 | 10 | 1,924 | 0.0052 |
| 6 | Latino2 | 3 | 1,068 | 0.0027 |
| 7 | MiddleEastern2 | 1 | 669 | 0.0016 |
| 8 | SouthAsian1 | 2 | 496 | 0.004 |
| 9 | EastAsian1 | 1 | 457 | 0.002 |
| 10 | European4 | 1 | 265 | 0.0028 |
| 11 | Admixed1 | 0 | 106 | 0 |
| Total |  | 172 | 22,530 | 0.0076 |

**Supplemental Table 6.**
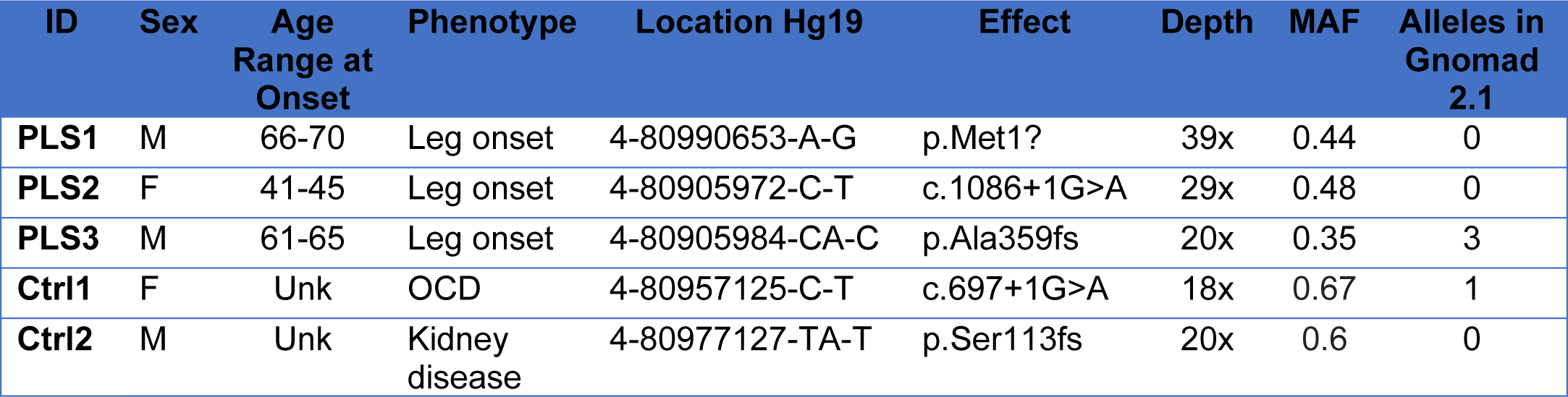
Description of PLS participants with *ANTXR2* PTV Qualifying Variants.

